# Recombinant adjuvanted zoster vaccine and reduced risk of COVID-19 diagnosis and hospitalization in older adults

**DOI:** 10.1101/2021.10.01.21264400

**Authors:** Katia J. Bruxvoort, Bradley Ackerson, Lina S. Sy, Amit Bhavsar, Hung Fu Tseng, Ana Florea, Yi Luo, Yun Tian, Zendi Solano, Robyn Widenmaier, Meng Shi, Robbert Van Der Most, Johannes Eberhard Schmidt, Jasur Danier, Thomas Breuer, Lei Qian

**Affiliations:** Department of Research & Evaluation, Kaiser Permanente Southern California, 100 S Los Robles, Pasadena, CA 91101, USA; Department of Epidemiology, University of Alabama at Birmingham, 1665 University Blvd, Birmingham, AL 35233, USA; GSK, 20 Avenue Fleming, 1300 Wavre, Belgium; Kaiser Permanente Bernard J. Tyson School of Medicine, 98 S. Los Robles Ave., Pasadena, CA 91101, USA; GSK, 14200 Shady Grove Road, Rockville, MD 20850, USA; GSK, 89 Rue de l’Institut, 1330 Rixensart, Belgium; GSK, 1 Via Fiorentina, 53100 Siena, Italy

**Keywords:** COVID-19, zoster vaccine, trained immunity, non-specific effects

## Abstract

**Background:** Vaccines may elicit long-term boosting of innate immune responses that can help protect against COVID-19. We evaluated the association between recombinant adjuvanted zoster vaccine (RZV) and COVID-19 outcomes at Kaiser Permanente Southern California.

**Methods:** In a cohort design, adults aged ≥50 years who received ≥1 RZV dose prior to 3/1/2020 were matched 1:2 to unvaccinated individuals and followed until 12/31/2020. Adjusted hazard ratios (aHR) and 95% confidence intervals (CIs) for COVID-19 outcomes were estimated using Cox proportional hazards regression. In a test-negative design, cases had a positive SARS-CoV-2 test and controls had only negative tests, from 3/1/2020-12/31/2020. Adjusted odds ratios (aOR) and 95% CIs for prior receipt of RZV were estimated using logistic regression.

**Results:** In the cohort design, 149,244 RZV recipients were matched to 298,488 unvaccinated individuals. The aHRs (95% CI) for COVID-19 diagnosis and hospitalization were 0.84 (0.81-0.87) and 0.68 (0.64-0.74), respectively. In the test-negative design, 8.4% of 75,726 test-positive cases and 13.1% of 340,898 test-negative controls had received ≥1 RZV dose. The aOR (95% CI) was 0.84 (0.81-0.86).

**Conclusion:** RZV vaccination was associated with a 16% lower risk of COVID-19 diagnosis and 32% lower risk of hospitalization, suggesting RZV elicits heterologous protection, possibly through trained immunity.

## Background

The severe acute respiratory syndrome coronavirus 2 (SARS-CoV-2) that causes coronavirus disease 2019 (COVID-19) has triggered a global pandemic with over 224 million infections and more than 4.6 million deaths.[1] Despite the extraordinary pace of COVID-19 vaccine development, nearly 12 months were required to develop, evaluate, and initiate large-scale production and administration of highly effective vaccines.[2] As of September 2021, COVID-19 vaccines have been available for over 9 months, but nearly 58% of the global population remains unvaccinated, despite ongoing efforts to increase global vaccine production and distribution.[3]

Traditionally, immune memory consisting of pathogen-specific cellular and humoral responses was thought to be the hallmark of the adaptive response. However, a growing body of evidence suggests that the innate immune system can also develop and maintain immune memory (i.e. trained immunity), which may be helpful in reducing the impact of a broad array of infectious diseases.[4-6] Several studies with vaccines, including the tuberculosis vaccine Bacillus Calmette-Guérin (BCG), and measles, oral polio, and influenza vaccines, including vaccine adjuvants, demonstrate the ability of the innate immune system to form memory and to provide non-specific protection against heterologous infections.[5-9]

Recombinant adjuvanted zoster vaccine (RZV) is the first approved and widely used vaccine with the novel AS01 adjuvant, which elicits an innate immune response and robust cellular and humoral responses.[10, 11] We hypothesized that RZV could induce trained immunity that might reduce SARS-CoV-2 infections in older adults, who are at high risk of severe disease and poor outcomes following SARS-CoV-2 infection.[12]

Therefore, we evaluated the association of receipt of RZV with COVID-19 diagnosis and COVID-19-associated hospitalizations in a large cohort of adults aged ≥50 years.

## Methods

### Study setting

We employed both matched cohort and test-negative designs in an observational study conducted at Kaiser Permanente Southern California (KPSC), an integrated health care system with 15 hospitals and 235 associated medical office buildings. The KPSC population of over 4.6 million members is stable and mirrors the sociodemographic, racial, and ethnic diversity of Southern California.[13] KPSC members are incentivized to seek care within the KPSC system, and recommended vaccinations are proactively offered free of charge at any visit or at walk-in nurse clinic visits. Comprehensive electronic health records (EHR) capture all details of patient care, including diagnoses, vaccinations, procedures, laboratory tests, and pharmacy records. Care received outside of KPSC is captured through claims.

Molecular diagnostic testing for SARS-CoV-2 evolved during the study period (3/1/2020-12/31/2020). In the early months of the pandemic, testing was prioritized for individuals with symptoms and prior to hospital admissions or certain outpatient procedures. RT-PCR was primarily conducted on nasopharyngeal/oropharyngeal swabs using the Roche cobas^®^ SARS-CoV-2 assay on the Roche cobas^®^ 6800 and 8800 analyzers (Roche Molecular Systems, New Jersey, USA) or the Aptima^®^ SARS-CoV-2 assay on Hologic Panther^®^ analyzers (Hologic Inc, San Diego, California, USA). However, testing guidelines were gradually expanded, such that by September 2020 testing was widely available for symptomatic and asymptomatic individuals for any reason. By November 2020, implementation of saliva testing and a new COVID-19 laboratory using the TaqPath™ COVID-19 High-Throughput Combo Kit on the Thermo Fisher Scientific Amplitude Solution (Thermo Fisher Scientific, California, USA) increased testing capacity at KPSC to approximately 52,000 tests per day.

### Cohort design

The cohort design included individuals aged ≥50 years as of 3/1/2020 who had at least 1-year prior KPSC membership (allowing for a 31-day gap). Separate analyses were conducted defining the exposure as either receipt of at least 1 dose of RZV prior to 3/1/2020, receipt of 2 doses of RZV ≥4 weeks apart prior to 3/1/2020, or receipt of 1 dose of RZV only prior to 3/1/2020. Outcomes were COVID-19 diagnosis, defined as a positive SARS-CoV-2 molecular test or COVID-19 diagnosis code (**Supplementary Table 1**), and COVID-19 hospitalization, defined as a hospitalization with a SARS-CoV-2 positive test or a COVID-19 diagnosis, or a hospitalization occurring within 7 days after a SARS-CoV-2 positive test. We excluded individuals with COVID-19 outcomes occurring ≤14 days after receipt of RZV.

Recipients of at least 1 dose of RZV as of 3/1/2020 were matched 1:2 with RZV unvaccinated individuals by age (50-59 years, 60-69 years, 70-79 years, and ≥80 years), sex, race/ethnicity (non-Hispanic White, non-Hispanic Black, Hispanic, non-Hispanic Asian, and other/unknown), and zip code. Individuals were followed through EHR from 3/1/2020 until occurrence of COVID-19 outcomes, termination of membership, death, receipt of a first dose of RZV for unvaccinated individuals, or end of the study period (12/31/2020), whichever came first.

We identified clinical characteristics in the year prior to 3/1/2020 including: most recent body mass index (BMI), most recent smoking status, health care utilization (number of outpatient visits, number of emergency department [ED] visits, number of hospitalizations), frailty using the algorithm described by Kim et al.,[14] comorbidities (cardiovascular disease, diabetes, hypertension, pulmonary disease, renal disease, cancer, autoimmune disease using *International Classification of Diseases Tenth Revision (ICD-10)* codes,[15] and HIV using the KPSC HIV registry), other vaccinations (influenza, pneumococcal, and tetanus, diphtheria, and acellular pertussis [Tdap]), and medical center area.

We used similar methods for the analysis comparing recipients of at least 1 dose of RZV and their unvaccinated matches, the analysis comparing recipients of 2 doses of RZV and their unvaccinated matches, and the analysis comparing recipients of 1 dose of RZV only and their unvaccinated matches. We first described characteristics of RZV vaccinated and RZV unvaccinated individuals, reporting absolute standardized differences (ASD) to assess balance between covariates; characteristics that were substantially different between RZV vaccinated and unvaccinated groups (ASD >0.1) were considered potential confounders for inclusion in multivariable analyses. We calculated incidence rates for COVID-19 outcomes by dividing the number of COVID-19 outcomes by the total number of person-years, and we used the Kaplan-Meier method to estimate cumulative incidence. Finally, we used Cox proportional hazards regression to estimate hazard ratios (HR) and 95% confidence intervals (CI) for COVID-19 outcomes comparing RZV vaccinated and unvaccinated individuals, adjusting for potential confounders. To further control for differences in health care seeking behavior by vaccination status and potential effect modification due to receipt of other vaccines, sensitivity analyses were conducted to compare the risk of COVID-19 outcomes among RZV vaccinated and unvaccinated individuals, in a subset of individuals who had received influenza vaccine but no other vaccines in the year prior to 3/1/2020. Analyses comparing recipients of at least 1 dose of RZV and their unvaccinated matches were performed among this subset of individuals.

### Test-negative design

The test-negative design included all individuals tested for SARS-CoV-2 from 3/1/2020 to 12/31/2020, who had at least 1-year prior membership (allowing for a 31-day gap) and were aged ≥50 years as of the SARS-CoV-2 test date. Test-positive cases comprised individuals with a positive SARS-CoV-2 test, and test-negative controls comprised those with only negative SARS-CoV-2 tests during the same period. For individuals with multiple SARS-CoV-2 tests, the test-positive case was defined as the first positive test for individuals with any positive tests, and the test-negative control was defined as the first negative test for individuals with only negative tests. Separate analyses were conducted defining the exposure as either receipt of at least 1 dose of RZV ≥14 days prior to the SARS-CoV-2 test or receipt of 2 doses of RZV ≥4 weeks apart ≥14 days prior to the SARS-CoV-2 test. Individuals who received RZV <14 days prior to the SARS-CoV-2 test were excluded from the study.

We described characteristics of test-positive cases and test-negative controls in the year prior to their SARS-CoV-2 test date, using similar methods as for the cohort design. We used logistic regression to estimate odds ratios (OR) and 95% CI comparing odds of RZV vaccination among test-positive cases and test-negative controls, adjusting for age, sex, race/ethnicity, calendar time, and other confounders based on ASD >0.1. We also conducted analyses stratifying the RZV exposure by time from most recent RZV dose to SARS-CoV-2 test (15 days to <1 month, 1 to <6 months, 6 months to <1 year, and ≥1 year).

The KPSC Institutional Review Board reviewed and approved the study with a waiver for the requirement of informed consent.

## Results

### Cohort design – receipt of at least 1 dose of RZV

The cohort design with at least 1 dose of RZV as the exposure included 149,244 RZV vaccinated individuals and 298,488 matched RZV unvaccinated individuals (**Table 1**). Overall, 16.2% were aged 50-59 years and 12.8% were ≥80 years, 57.8% were female, and 54.1% were non-Hispanic White. Recipients of at least 1 dose of RZV had less missing data on BMI (2.6% vs 13.2% of matched RZV unvaccinated individuals, ASD 0.41) and on smoking (2.7% vs 12.7%, ASD 0.39), and had more outpatient visits in the year prior to 3/1/2020 (39.1% vs 28.8% with ≥11 visits, ASD 0.49). They more commonly had hypertension (49.0% vs 43.9%, ASD 0.10) and had received other vaccinations (93.6% vs 73.3%, ASD 0.57) in the year prior. Other characteristics, including the number of ED visits and hospitalizations, frailty, other comorbidities in the year prior, and medical center area, were well-balanced between recipients of at least 1 dose of RZV and their RZV unvaccinated matches.

**Table 1.** Baseline characteristics of RZV (at least 1 dose) vaccinated and unvaccinated cohort.

|  | Vaccinated<br>(N=149244)<br>n (%) | Unvaccinated<br>(N=298488)<br>n (%) | Absolute<br>Standardized<br>Difference<sup>d</sup> |
| --- | --- | --- | --- |
| Age at index date, years |  |  | N/A<sup>c</sup> |
| 50-59 | 24169 (16.2) | 48338 (16.2) |  |
| 60-69 | 56047 (37.6) | 112094 (37.6) |  |
| 70-79 | 49986 (33.5) | 99972 (33.5) |  |
| ≥80 | 19042 (12.8) | 38084 (12.8) |  |
| Sex |  |  | N/A<sup>c</sup> |
| Female | 86206 (57.8) | 172412 (57.8) |  |
| Male | 63038 (42.2) | 126076 (42.2) |  |
| Race/Ethnicity |  |  | N/A<sup>c</sup> |
| Non-Hispanic White | 80743 (54.1) | 161486 (54.1) |  |
| Non-Hispanic Black | 8411 (5.6) | 16822 (5.6) |  |
| Hispanic | 30376 (20.4) | 60752 (20.4) |  |
| Non-Hispanic Asian | 24434 (16.4) | 48868 (16.4) |  |
| Other/Unknown | 5280 (3.5) | 10560 (3.5) |  |
| Body mass index<sup>a</sup> |  |  | 0.41 |
| <18.5 | 1925 (1.3) | 4418 (1.5) |  |
| 18.5-<25 | 45482 (30.5) | 76156 (25.5) |  |
| 25-<30 | 54387 (36.4) | 93932 (31.5) |  |
| 30-<35 | 27736 (18.6) | 52216 (17.5) |  |
| 35-<40 | 10398 (7.0) | 20911 (7.0) |  |
| 40-<45 | 3647 (2.4) | 7511 (2.5) |  |
| ≥45 | 1734 (1.2) | 3993 (1.3) |  |
| Unknown | 3935 (2.6) | 39351 (13.2) |  |
| Smoking<sup>a</sup> |  |  | 0.39 |
| No | 111269 (74.6) | 194333 (65.1) |  |
| Yes | 34016 (22.8) | 66278 (22.2) |  |
| Unknown | 3959 (2.7) | 37877 (12.7) |  |
| Number of outpatient visits<sup>b</sup> |  |  | 0.49 |
| 0 | 862 (0.6) | 26145 (8.8) |  |
| 1-4 | 33628 (22.5) | 95907 (32.1) |  |
| 5-10 | 56401 (37.8) | 90503 (30.3) |  |
| ≥11 | 58353 (39.1) | 85933 (28.8) |  |
| Number of emergency department visits<sup>b</sup> |  |  | 0.05 |
| 0 | 121729 (81.6) | 238943 (80.1) |  |
| 1 | 19135 (12.8) | 39466 (13.2) |  |
| ≥2 | 8380 (5.6) | 20079 (6.7) |  |
| Number of hospitalizations<sup>b</sup> |  |  | 0.01 |
| 0 | 130826 (87.7) | 261940 (87.8) |  |
| 1 | 11291 (7.6) | 22608 (7.6) |  |
| ≥2 | 7127 (4.8) | 13940 (4.7) |  |
| Frailty (top quartile)<sup>b</sup> | 37951 (25.4) | 73969 (24.8) | 0.01 |
| Baseline comorbidities<sup>b</sup> |  |  |  |
| Cardiovascular disease | 45251 (30.3) | 81069 (27.2) | 0.07 |
| Diabetes | 34703 (23.3) | 69237 (23.2) | 0.00 |
| Hypertension | 73201 (49.0) | 131033 (43.9) | 0.10 |
| Pulmonary disease | 23628 (15.8) | 41768 (14.0) | 0.05 |
| Renal disease | 18965 (12.7) | 37487 (12.6) | 0.00 |
| Cancer | 8484 (5.7) | 17322 (5.8) | 0.01 |
| HIV | 1201 (0.8) | 818 (0.3) | 0.07 |
| Autoimmune disease | 7566 (5.1) | 13681 (4.6) | 0.02 |
| Other vaccinations<sup>b</sup> | 139723 (93.6) | 218851 (73.3) | 0.57 |
| Influenza vaccine | 136984 (98.0) | 211906 (96.8) |  |
| PCV13/PPSV23 | 17920 (12.8) | 25737 (11.8) |  |
| Tdap | 13806 (9.9) | 17254 (7.9) |  |

Abbreviations: PCV13, 13-valent pneumococcal conjugate vaccine; PPSV23, pneumococcal polysaccharide vaccine; RZV, recombinant zoster vaccine; Tdap, tetanus, diphtheria, acellular pertussis vaccine
<sup>a</sup> Most recent in 365 days prior to 3/1/2020
<sup>b</sup> In 365 days prior to 3/1/2020
<sup>c</sup> N/A= not applicable, for matching variable
<sup>d</sup> Potential confounders were determined by absolute standardized difference >0.1
Medical center area not shown. There were no significant differences in the distribution of the vaccinated and unvaccinated individuals across the 19 medical center areas.

Among recipients of at least 1 dose of RZV, there were 5,951 COVID-19 diagnoses and 1,066 COVID-19 hospitalizations, with incidence rates per 1,000 person-years (95% CI) of 48.82 (47.60-50.08) and 8.69 (8.18-9.23), respectively (**Table 2**). Among RZV unvaccinated individuals matched to recipients of at least 1 dose of RZV, there were 13,028 COVID-19 diagnoses and 2,765 COVID-19 hospitalizations, with incidence rates per 1,000 person-years of 55.01 (54.07-55.96) and 11.59 (11.17-12.03), respectively. In Kaplan-Meier analyses, the cumulative incidences of COVID-19 diagnosis and COVID-19 hospitalization were lower among recipients of at least 1 dose of RZV compared to their RZV unvaccinated matches (**Figure 1-2**). In fully adjusted analyses, recipients of at least 1 dose of RZV had a 16% lower rate of COVID-19 diagnosis (HR 0.84 [95% CI: 0.81-0.87]) and a 32% lower rate of COVID-19 hospitalization (HR 0.68 [95% CI: 0.64-0.74]) compared to RZV unvaccinated individuals (**Table 2**).

**Figure 1:**
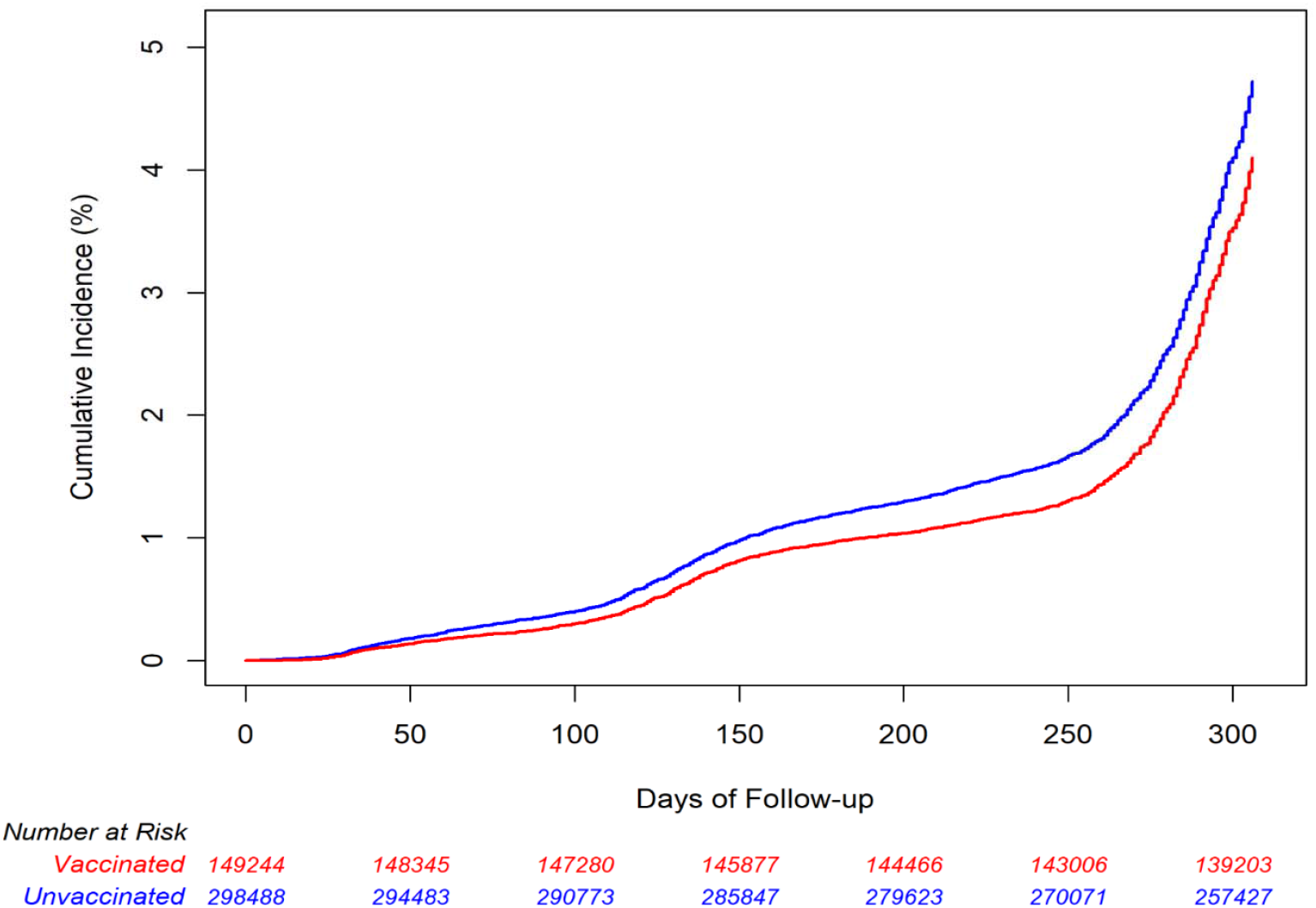
Cumulative incidence estimates of COVID-19 diagnosis by RZV (at least 1 dose) vaccination status. Abbreviations: COVID-19, coronavirus disease 2019; RZV, recombinant zoster vaccine

**Figure 2:**
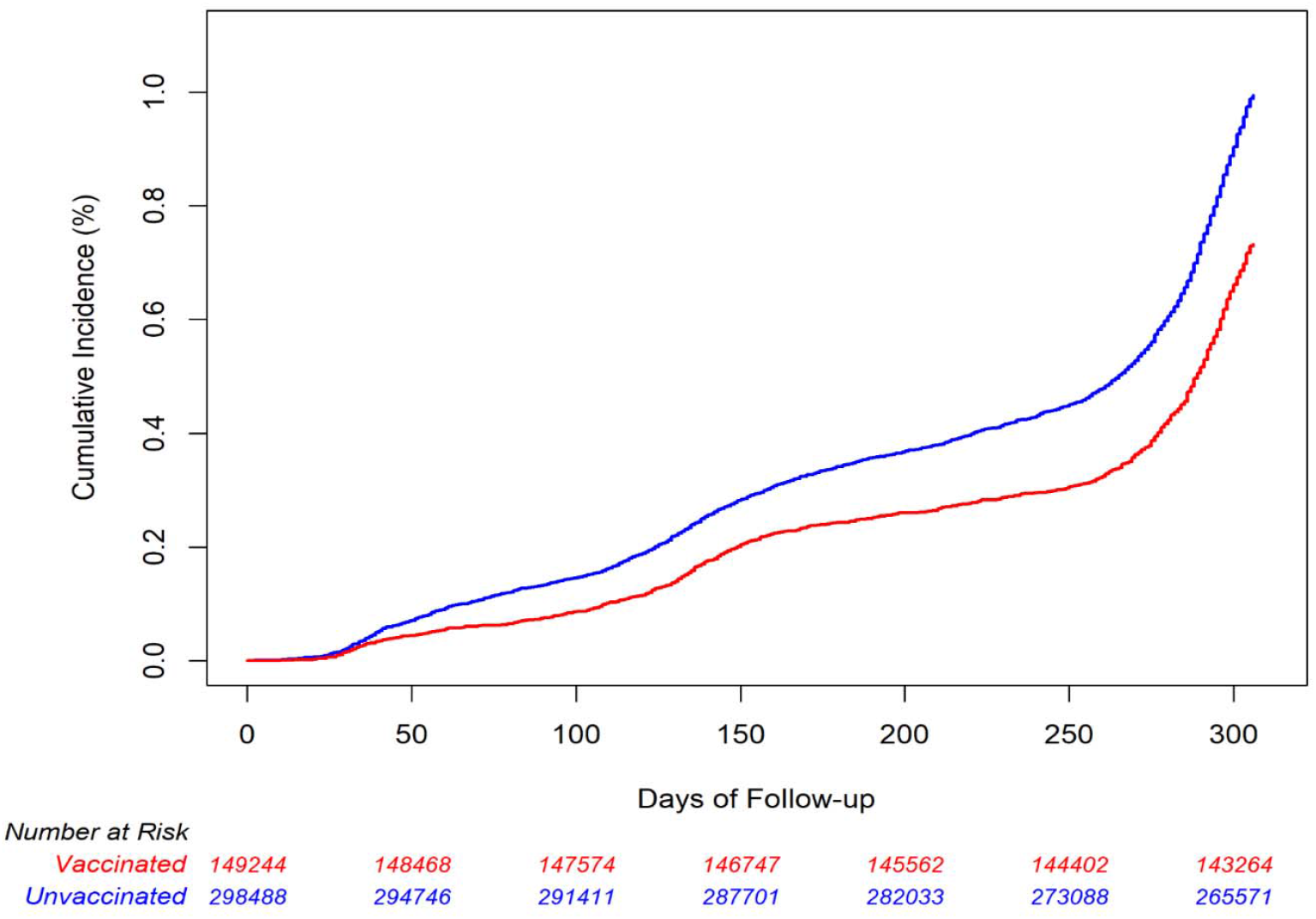
Cumulative incidence estimates of COVID-19 hospitalization by RZV (at least 1 dose) vaccination status. Abbreviations: COVID-19, coronavirus disease 2019; RZV, recombinant zoster vaccine

**Table 2.** Incidence rates and hazard ratios of COVID-19 diagnosis and hospitalization among vaccinated (at least 1 dose of RZV) versus unvaccinated individuals.

| Outcomes | Vaccinated<br>(N=149244) |  |  | Unvaccinated<br>(N=298488) |  |  | Hazard Ratio (95% CI) |  |
| --- | --- | --- | --- | --- | --- | --- | --- | --- |
|  | Number<br>of cases | Number of<br>person-years | Incidence per 1000<br>person-years<br>(95% CI) | Number<br>of cases | Number of<br>person-years | Incidence per 1000<br>person-years<br>(95% CI) | Unadjusted | Adjusted<sup>a</sup> |
| COVID-19 diagnosis | 5951 | 121887.27 | 48.82 (47.60-50.08) | 13028 | 236826.63 | 55.01 (54.07-55.96) | 0.86 (0.84-0.89) | 0.84 (0.81-0.87) |
| COVID-19 hospitalization | 1066 | 122689.85 | 8.69 (8.18-9.23) | 2765 | 238530.10 | 11.59 (11.17-12.03) | 0.73 (0.68-0.79) | 0.68 (0.64-0.74) |

Abbreviations: COVID-19, coronavirus disease 2019; RZV, recombinant zoster vaccine
<sup>a</sup> Adjusted for covariates: body mass index, smoking, number of outpatient visits, hypertension, and other vaccinations.

In sensitivity analyses among the subset of individuals who had received influenza vaccine but no other vaccines in the year prior to 3/1/2020, we observed similar reductions in COVID-19 diagnosis and hospitalization among recipients of at least 1 dose of RZV compared to RZV unvaccinated individuals (**Supplementary Table 2)**. In fully adjusted analyses of this subset, recipients of at least 1 dose of RZV had a 17% lower rate of COVID-19 diagnosis (HR 0.83 [95% CI: 0.78-0.89]) and a 32% lower rate of COVID-19 hospitalization (HR 0.68 [95% CI: 0.59-0.78]).

### Cohort design – receipt of 2 doses of RZV

The cohort design with 2 doses of RZV as the exposure included 94,895 RZV vaccinated individuals and 189,790 matched RZV unvaccinated individuals (**Supplementary Table 3**). Differences in characteristics among recipients of 2 doses of RZV and their RZV unvaccinated matches were similar to the distributions observed for recipients of at least 1 dose of RZV and their RZV unvaccinated matches.

Among recipients of 2 doses of RZV, there were 3,403 COVID-19 diagnoses and 612 COVID-19 hospitalizations, with incidence rates per 1,000 person-years (95% CI) of 43.79 (42.34-45.28) and 7.83 (7.23-8.48), respectively (**Table 3**). Among unvaccinated individuals matched to recipients of 2 doses of RZV, there were 7,689 COVID-19 diagnoses and 1,676 COVID-19 hospitalizations, with incidence rates per 1,000 person-years of 51.03 (49.90-52.19) and 11.05 (10.53-11.59), respectively. In Kaplan-Meier analyses, the cumulative incidences of COVID-19 diagnosis and COVID-19 hospitalization were lower among recipients of 2 doses of RZV compared to their RZV unvaccinated matches (**Supplementary Figures 1-2**). In fully adjusted analyses, those who received 2 doses of RZV had a 19% lower rate of COVID-19 diagnosis (HR 0.81 [95% CI: 0.77-0.84]) and a 36% lower rate of COVID-19 hospitalization (HR 0.64 [95% CI: 0.58-0.70]) (**Table 3**).

**Table 3.** Incidence rates and hazard ratios of COVID-19 diagnosis and hospitalization among vaccinated (2 doses of RZV) versus unvaccinated individuals.

| Outcomes | Vaccinated<br>(N=94895) |  |  | Unvaccinated<br>(N=189790) |  |  | Hazard Ratio (95% CI) |  |
| --- | --- | --- | --- | --- | --- | --- | --- | --- |
|  | Number<br>of cases | Number of<br>person-years | Incidence per 1000<br>person-years<br>(95% CI) | Number<br>of cases | Number of<br>person-years | Incidence per 1000<br>person-years<br>(95% CI) | Unadjusted | Adjusted<sup>a</sup> |
| COVID-19 diagnosis | 3403 | 77714.97 | 43.79 (42.34-45.28) | 7689 | 150668.58 | 51.03 (49.90-52.19) | 0.83 (0.80-0.87) | 0.81 (0.77-0.84) |
| COVID-19 hospitalization | 612 | 78164.05 | 7.83 (7.23-8.48) | 1676 | 151656.50 | 11.05 (10.53-11.59) | 0.69 (0.63-0.76) | 0.64 (0.58-0.70) |

Abbreviations: COVID-19, coronavirus disease 2019; RZV, recombinant zoster vaccine
<sup>a</sup> Adjusted for covariates: body mass index, smoking, number of outpatient visits, and other vaccinations.

### Cohort design – receipt of 1 dose of RZV only

The cohort design with 1 dose of RZV only as the exposure included 54,349 RZV vaccinated individuals and 108,698 matched RZV unvaccinated individuals (**Supplementary Table 4**). Results of this analysis were similar, but less pronounced, than results of the analyses for the other exposures (receipt of at least 1 dose of RZV and receipt of 2 doses of RZV). Recipients of 1 dose of RZV only had a 14% lower rate of COVID-19 diagnosis (HR 0.86 [95% CI: 0.82-0.91]) and 24% lower rate of COVID-19 hospitalization (HR 0.76 [95% CI: 0.68-0.86]).

### Test-negative design

The test-negative design included 75,726 test-positive COVID-19 cases and 340,898 test-negative controls (**Table 4**). Cases were younger than controls (49.0% vs 36.1% ages 50-59 years, ASD 0.31) and less often non-Hispanic White (25.2% vs 43.4%, ASD 0.50). A higher proportion of cases were obese, and a lower proportion of cases were smokers as compared to controls, but cases also had more missing data for these variables. Cases also had fewer outpatient and ED visits in the year prior to the SARS-CoV-2 test date than controls (16.3% vs 27.8% with ≥11 outpatient visits, ASD 0.36, and 6.5% vs 9.4% with ≥2 ED visits, ASD 0.14), were less frail (17.8% vs 26.6% in the top quartile of the frailty index, ASD 0.21), less commonly had chronic comorbidities (cardiovascular disease, hypertension, pulmonary disease, renal disease, and cancer), and less commonly had received other vaccinations in the year prior (68.3% vs 76.7%, ASD 0.19). There were also significant differences between cases and controls in test month and medical center area.

**Table 4.**
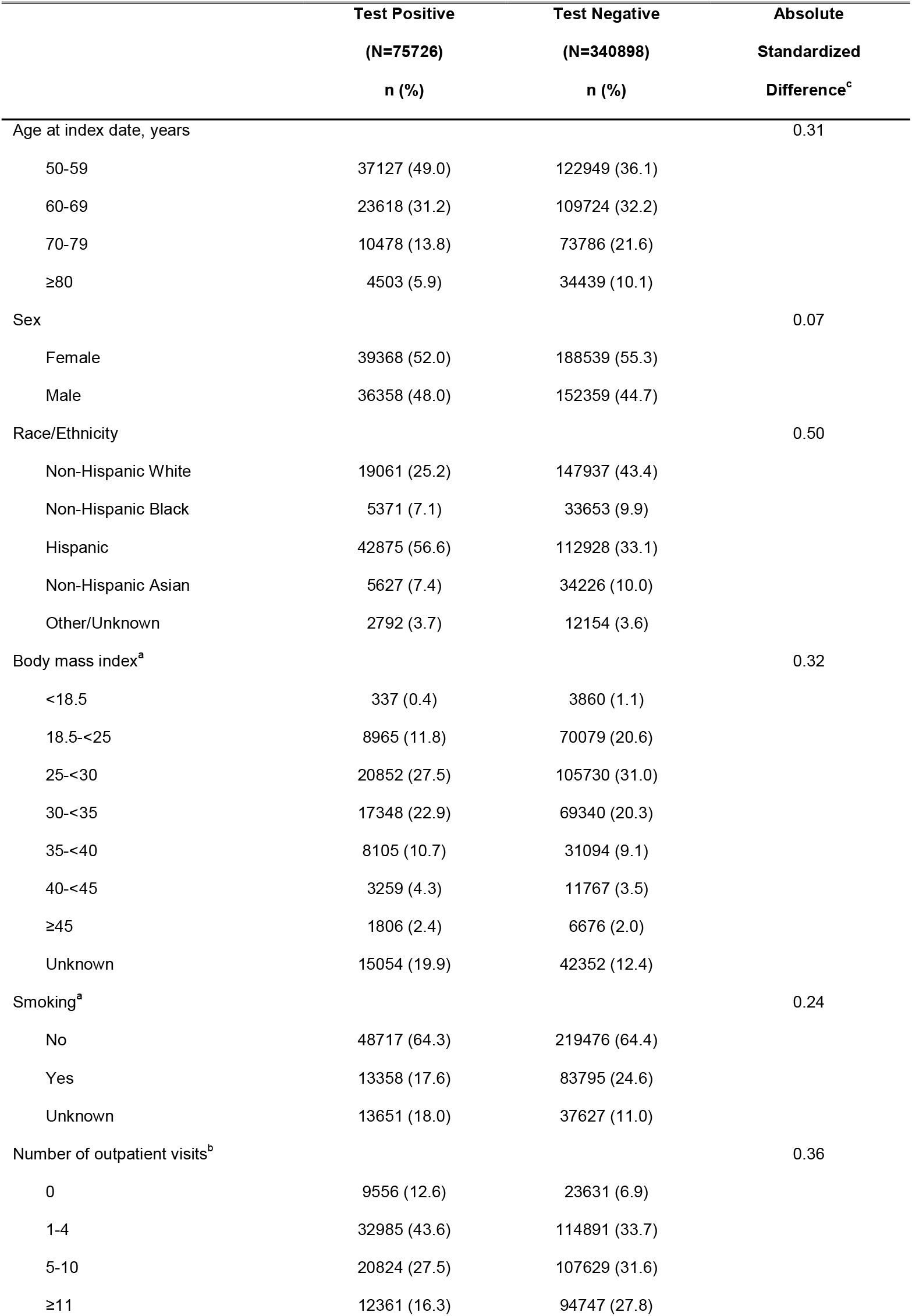
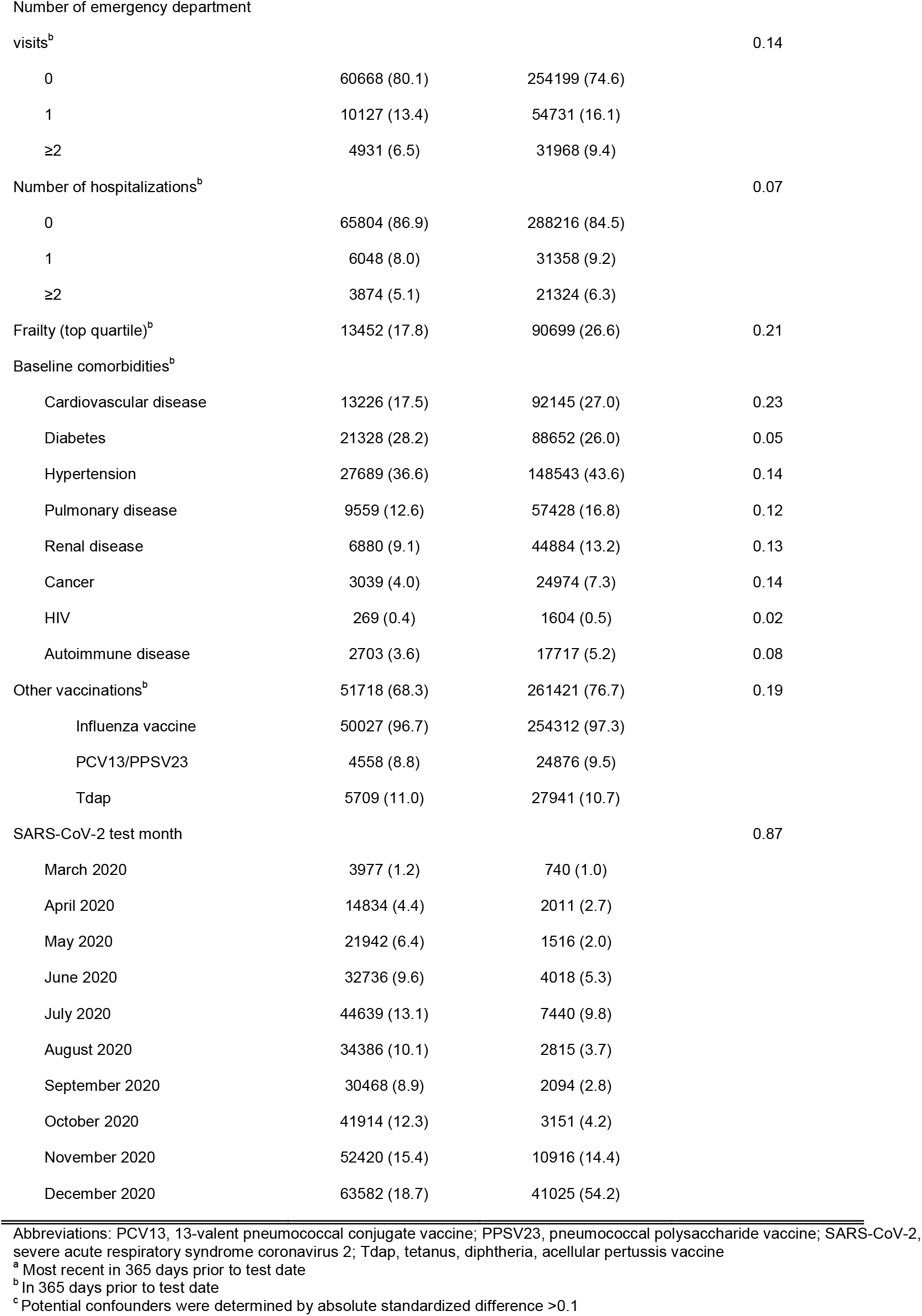
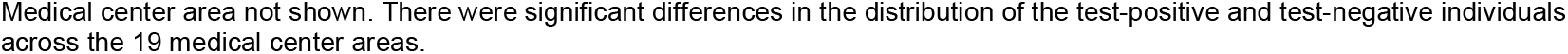
Characteristics of SARS-CoV-2 test-positive cases and test-negative controls.

Of cases and controls, respectively, 8.4% and 13.1% had received at least 1 dose of RZV, 5.4% and 9.2% had received 2 doses of RZV, and 91.6% and 86.9% were RZV unvaccinated (**Table 5**). The adjusted ORs (95% CI) comparing cases and controls were 0.84 (0.81-0.86) for individuals who received at least 1 dose of RZV vs unvaccinated individuals and 0.82 (0.79-0.85) for individuals who received 2 doses of RZV vs unvaccinated individuals. Adjusted ORs did not appear to vary substantially by time since RZV vaccination.

**Table 5.** Odds ratio of RZV vaccination among SARS-CoV-2 test-positive cases versus test-negative controls.

| Exposure | Test Positive |  | Test Negative |  | Odds Ratio (95% CI) |  |
| --- | --- | --- | --- | --- | --- | --- |
|  | (N=75726) |  | (N=340898) |  |  |  |
|  | n | % | n | % | Unadjusted | Adjusted<sup>a</sup> |
| RZV (at least 1 dose) vaccinated | 6392 | 8.4 | 44786 | 13.1 | 0.61 (0.59-0.63) | 0.84 (0.81-0.86) |
| 15 days to <1 month<sup>b</sup> | 302 | 0.4 | 1810 | 0.5 | 0.71 (0.63-0.81) | 0.79 (0.69-0.90) |
| 1 to <6 months | 1703 | 2.3 | 10643 | 3.1 | 0.68 (0.65-0.72) | 0.87 (0.82-0.92) |
| 6 months to <1 year | 1697 | 2.2 | 14606 | 4.3 | 0.50 (0.47-0.52) | 0.83 (0.78-0.87) |
| ≥1 year | 2690 | 3.6 | 17727 | 5.2 | 0.65 (0.62-0.68) | 0.82 (0.79-0.86) |
| RZV (2 doses) vaccinated | 4108 | 5.4 | 31359 | 9.2 | 0.56 (0.54-0.58) | 0.82 (0.79-0.85) |
| RZV unvaccinated | 69334 | 91.6 | 296112 | 86.9 | N/A<sup>c</sup> | N/A<sup>c</sup> |

Abbreviation: RZV, recombinant zoster vaccine; SARS-CoV-2, severe acute respiratory syndrome coronavirus 2
<sup>a</sup> Adjusted for covariates age, sex, race/ethnicity, calendar time, body mass index, smoking, number of outpatient visits, number of emergency department visits, frailty, cardiovascular disease, hypertension, pulmonary disease, renal disease, cancer, other vaccinations, and medical center area.
<sup>b</sup> Time since most recent RZV vaccination
<sup>c</sup> N/A= not applicable

## Discussion

This large retrospective study spanning the first year of the COVID-19 pandemic provides evidence from both cohort and test-negative designs that receipt of RZV prior to the availability of COVID-19 vaccines may have provided benefit in reducing the burden of COVID-19 in adults aged ≥50 years. In the cohort design analysis, after adjusting for potential confounders including other vaccinations, we found that the risk of COVID-19 diagnosis was reduced by 16% among persons who had received at least 1 dose of RZV compared to RZV unvaccinated individuals, an association that was similar and did not vary by time since most recent RZV dose in the test-negative analysis. We also found that the risk of hospitalization with COVID-19 was reduced by 32% among recipients of at least 1 dose of RZV compared to RZV unvaccinated individuals. These results were consistent in the test-negative design (16% lower odds of RZV vaccination among SARS-CoV-2 test-positive cases versus test-negative controls).

A similar reduction in risk of COVID-19 infection and severe disease has been reported following receipt of influenza vaccine in several studies [16-18], although this association was not observed in other studies.[19-23] A prior study at KPSC found a reduction in risk of COVID-19 diagnosis and severe disease associated with receipt of 13-valent pneumococcal conjugate vaccine (PCV13) but not with receipt of influenza vaccine.[24] Interestingly, in a sensitivity analysis among the subset of individuals who received influenza vaccine but no other vaccines, we found that receipt of RZV was still associated with a similar reduction in risk of COVID-19 diagnosis and hospitalization, suggesting that influenza vaccination or healthy vaccinee bias had minimal impact on our findings.

While the mechanism for the reduced risk of heterologous infections following receipt of some vaccines is not clear, it is plausible that these vaccines induce a trained immune response that results in an improved cytokine response to subsequent exposures; this response may provide an antiviral effect against infection with SARS-CoV-2 or other viruses.[25] Induction of an innate immune response has been suggested as a means to control viral replication early in the course of infection to reduce the risk of severe disease.[4, 9, 25] This may be particularly important for viral infections such as SARS-CoV-2, which appear to attenuate the host innate immune response as a means of increasing viral replication, possibly increasing disease severity and enhancing viral transmission.[26, 27] Notably, induction of innate immunity by highly-effective COVID-19 vaccines has been proposed as a possible mechanism of early protection following vaccination prior to the development of a robust adaptive response.[28] Furthermore, innate immunity has been found to play an important role in the control of infection in coronavirus animal reservoirs and may similarly contribute to reduced severity of COVID-19 disease observed in children compared to adults.[29, 30]

Our study found a durable reduction in the risk of COVID-19 infection following receipt of RZV vaccine, consistent with the durable protection against heterologous infections provided by trained immunity.[9] It is possible that the AS01 adjuvant used in RZV, which activates innate immune responses including monocytes and the IFN-response pathways, may be associated with the reduced risk of COVID-19 diagnosis and hospitalization observed in this study.[10, 11, 31] Influenza and other vaccines may also induce similar trained immunity against SARS-CoV-2 infection, although the duration may be variable.[9, 25, 32-34] Additional research is needed to elucidate immunologic mechanisms for our findings and to explore implications such as trained immunity-based vaccines that might mitigate serious infections in future pandemics until specific vaccines become available.[35]

Our study had several strengths and limitations. We leveraged comprehensive EHR data on demographic and clinical characteristics, vaccinations, and COVID-19 outcomes from a large, diverse cohort of adults aged ≥50 years. Although data for BMI and smoking status were missing for some individuals, we conducted a sensitivity analysis limited to individuals without missing data and found similar results. As in all observational studies, there may be some residual confounding. RZV recipients may have differed from RZV unvaccinated individuals with respect to health status, health care seeking behavior, and other factors that might, in part, explain the observed differences in risk of COVID-19 diagnosis and hospitalization. However, to reduce potential confounding, we used a cohort design, matching RZV recipients with unvaccinated individuals on age, sex, race/ethnicity, and zip code, and adjusting for health care utilization, other vaccinations, and comorbidities. We also used a test-negative design, which may be less confounded by health care seeking behavior than the cohort design,[36] but may also be less generalizable given that the study population was limited to those who were tested for SARS-CoV-2. We observed similar results using both designs, supporting the validity of the results. In addition, RZV exposure could be misclassified if individuals were vaccinated outside of KPSC; however, such misclassification was likely minimal, because KPSC members received vaccines at KPSC without charge, and providers were required to document previous receipt of recommended vaccines at all encounters.

## Conclusions

RZV recipients aged ≥50 years had a reduced risk of COVID-19 diagnosis and hospitalization compared to RZV unvaccinated individuals, suggesting that RZV may elicit durable innate immune responses that could offer heterologous protection against COVID-19 infection. While this epidemiological study provides further evidence of the concept of trained innate immunity using the adjuvanted RZV vaccine, additional research is needed to identify and understand the underlying non-specific effects of RZV and other vaccines.

## Supporting information

STROBE checklist

## Data Availability

The study details (GSK study number 217021) are available on the GSK Clinical Study Register and can be accessed at www.gskclinicalstudyregister.com.

https://www.gsk-studyregister.com/en/trial-details/?id=217021

## Acknowledgements

The authors thank the members of Kaiser Permanente for helping to improve care through the use of information collected through our electronic health record systems. Editorial and coordination support was provided by Adrian Kremer (Modis c/o GSK).

