## Supplemental materials for "Recombinant adjuvanted zoster vaccine and reduced risk of COVID-19 diagnosis and hospitalization in older adults"

**Supplementary Table 1. COVID-19 diagnosis codes**

| DXID[1] | Description | ICD-10 codes |
|---|---|---|
| 12459073 | CORONAVIRUS COVID-19 ACUTE RESPIRATORY DISTRESS SYNDROME | B97.29, J80, U07.1 |
| 12459074 | CORONAVIRUS COVID-19 PNEUMONIA | B97.29, J12.82, J12.89, U07.1 |
| 12459075 | CORONAVIRUS COVID-19 ACUTE BRONCHITIS | B97.29, J20.8, U07.1 |
| 12459076 | CORONAVIRUS COVID-19 LOWER RESPIRATORY INFECTION | B97.29, J22, U07.1 |
| 12459077 | ASYMPTOMATIC CORONAVIRUS COVID-19 DISEASE | B34.2, U07.1 |
| 12459078 | CORONAVIRUS COVID-19 DISEASE | B34.2, U07.1 |
| 12459108 | COVID-19 | U07.1 |
| 12459208 | CORONAVIRUS COVID-19 TEST POSITIVE BY OUTSIDE LABORATORY | U07.1 |
| 12460641 | CORONAVIRUS COVID-19 MULTISYSTEM INFLAMMATORY SYNDROME IN ADULT | M35.8, M35.81, U07.1 |
| 12460683 | PNEUMONIA DUE TO CORONAVIRUS DISEASE 2019 | J12.82 |

[1]KPSC uses internal codes termed “DXIDs” (lefthand column) for COVID-19 diagnoses, which each have a specific description for COVID-19 (middle column). The DXIDs correspond to ICD-10 codes (righthand column), which were not used in data analyses but are shown for reference.

**Supplementary Table 2. Incidence rates and hazard ratios of COVID-19 diagnosis and hospitalization among vaccinated (at least 1 dose of RZV) versus unvaccinated individuals, among a subset of individuals who received influenza vaccination but no other vaccinations in the year prior to 3/1/2020**

| | Vaccinated (N=37513) | | | Unvaccinated (N=75026) | | | Hazard Ratio (95% CI) | |
|---|---|---|---|---|---|---|---|---|
| **Outcomes** | **Number of cases** | **Number of person-years** | **Incidence per 1000 person-years (95% CI)** | **Number of cases** | **Number of person-years** | **Incidence per 1000 person-years (95% CI)** | **Unadjusted** | **Adjusted[a]** |
| COVID-19 diagnosis | 1356 | 30677.70 | 44.20 (41.91-46.62) | 3070 | 59409.98 | 51.67 (49.88-53.54) | 0.84 (0.79-0.90) | 0.83 (0.78-0.89) |
| COVID-19 hospitalization | 290 | 30850.43 | 9.40 (8.38-10.55) | 816 | 59765.63 | 13.65 (12.75-14.62) | 0.68 (0.59-0.78) | 0.68 (0.59-0.78) |

Abbreviations: COVID-19, coronavirus disease 2019; RZV, recombinant zoster vaccine
[a] Adjusted for covariates: body mass index, smoking, number of outpatient visits, and hypertension

**Supplementary Table 3. Baseline characteristics of RZV (2 doses) vaccinated and unvaccinated cohort**

| | Vaccinated<br>N = 94895<br>n (%) | Unvaccinated<br>N = 189790<br>n (%) | Absolute<br>Standardized Difference[d] |
|---|---|---|---|
| Age at index date, years | | | N/A[c] |
| 50-59 | 13102 (13.8) | 26204 (13.8) | |
| 60-69 | 35004 (36.9) | 70008 (36.9) | |
| 70-79 | 33823 (35.6) | 67646 (35.6) | |
| ≥80 | 12966 (13.7) | 25932 (13.7) | |
| Sex | | | N/A[c] |
| Female | 55755 (58.8) | 111510 (58.8) | |
| Male | 39140 (41.2) | 78280 (41.2) | |
| Race/Ethnicity | | | N/A[c] |
| Non-Hispanic White | 54371 (57.3) | 108742 (57.3) | |
| Non-Hispanic Black | 4324 (4.6) | 8648 (4.6) | |
| Hispanic | 16865 (17.8) | 33730 (17.8) | |
| Non-Hispanic Asian | 16057 (16.9) | 32114 (16.9) | |
| Other/Unknown | 3278 (3.5) | 6556 (3.5) | |
| Body mass index[a] | | | 0.41 |
| <18.5 | 1347 (1.4) | 2989 (1.6) | |
| 18.5-<25 | 30662 (32.3) | 50332 (26.5) | |
| 25-<30 | 34477 (36.3) | 59562 (31.4) | |
| 30-<35 | 16757 (17.7) | 32653 (17.2) | |
| 35-<40 | 6174 (6.5) | 12904 (6.8) | |
| 40-<45 | 2154 (2.3) | 4566 (2.4) | |
| ≥45 | 985 (1.0) | 2393 (1.3) | |
| Unknown | 2339 (2.5) | 24391 (12.9) | |
| Smoking[a] | | | 0.39 |
| No | 71024 (74.8) | 123695 (65.2) | |
| Yes | 21605 (22.8) | 42639 (22.5) | |
| Unknown | 2266 (2.4) | 23456 (12.4) | |
| Number of outpatient visits[b] | | | 0.56 |
| 0 | 329 (0.3) | 16035 (8.4) | |
| 1-4 | 17163 (18.1) | 59974 (31.6) | |
| 5-10 | 36790 (38.8) | 57975 (30.5) | |
| ≥11 | 40613 (42.8) | 55806 (29.4) | |
| Number of emergency department visits[b] | | | 0.06 |
| 0 | 77527 (81.7) | 151354 (79.7) | |
| 1 | 12122 (12.8) | 25455 (13.4) | |
| ≥2 | 5246 (5.5) | 12981 (6.8) | |
| Number of hospitalizations[b], | | | 0.01 |

| | | | |
|---|---|---|---|
| 0 | 83334 (87.8) | 166684 (87.8) | |
| 1 | 7053 (7.4) | 14395 (7.6) | |
| ≥2 | 4508 (4.8) | 8711 (4.6) | |
| Frailty (top quartile)[b] | 24050 (25.3) | 47113 (24.8) | 0.01 |
| Baseline comorbidities[b] | | | |
| Cardiovascular disease | 29971 (31.6) | 53747 (28.3) | 0.07 |
| Diabetes | 21254 (22.4) | 43645 (23.0) | 0.01 |
| Hypertension | 46781 (49.3) | 84396 (44.5) | 0.10 |
| Pulmonary disease | 15225 (16.0) | 26965 (14.2) | 0.05 |
| Renal disease | 12381 (13.0) | 24840 (13.1) | 0.00 |
| Cancer | 5590 (5.9) | 11372 (6.0) | 0.00 |
| HIV | 773 (0.8) | 493 (0.3) | 0.08 |
| Autoimmune disease | 4950 (5.2) | 8936 (4.7) | 0.02 |
| Other vaccinations[b] | 90080 (94.9) | 140588 (74.1) | 0.60 |
| Influenza vaccine | 88710 (98.5) | 136220 (96.9) | |
| PCV13/PPSV23 | 10774 (12.0) | 16442 (11.7) | |
| Tdap | 8375 (9.3) | 11071 (7.9) | |

Abbreviations: PCV13, 13-valent pneumococcal conjugate vaccine; PPSV23, pneumococcal polysaccharide vaccine; RZV, recombinant zoster vaccine; Tdap, tetanus, diphtheria, acellular pertussis vaccine
[a] Most recent in 365 days prior to 3/1/2020
[b] In 365 days prior to 3/1/2020
[c] N/A= not applicable, for matching variable
[d] Potential confounders were determined by absolute standardized difference >0.1
Medical center area not shown. There were no significant differences in the distribution of the vaccinated and unvaccinated individuals across the 19 medical center areas.

**Supplementary Table 4. Incidence rates and hazard ratios of COVID-19 diagnosis and hospitalization among vaccinated (1 dose of RZV only) versus unvaccinated individuals**

| Outcomes | Vaccinated (N=54349) | | | Unvaccinated (N=108698) | | | Hazard Ratio (95% CI) | |
|---|---|---|---|---|---|---|---|---|
| | Number of cases | Number of person-years | Incidence per 1000 person-years (95% CI) | Number of cases | Number of person-years | Incidence per 1000 person-years (95% CI) | Unadjusted | Adjusted[a] |
| COVID-19 diagnosis | 2548 | 44172.30 | 57.68 (55.49-59.97) | 5339 | 86158.05 | 61.97 (60.33-63.65) | 0.92 (0.88-0.97) | 0.86 (0.82-0.91) |
| COVID-19 hospitalization | 454 | 44525.79 | 10.20 (9.30-11.18) | 1089 | 86873.60 | 12.54 (11.81-13.30) | 0.81 (0.73-0.91) | 0.76 (0.68-0.86) |

Abbreviations: COVID-19, coronavirus disease 2019; RZV, recombinant zoster vaccine
[a] Adjusted for covariates: body mass index, smoking, number of outpatient visits, hypertension, and other vaccinations.

**Supplementary Figure 1: Cumulative incidence estimates of COVID-19 diagnosis by RZV (2 dose) vaccination status**

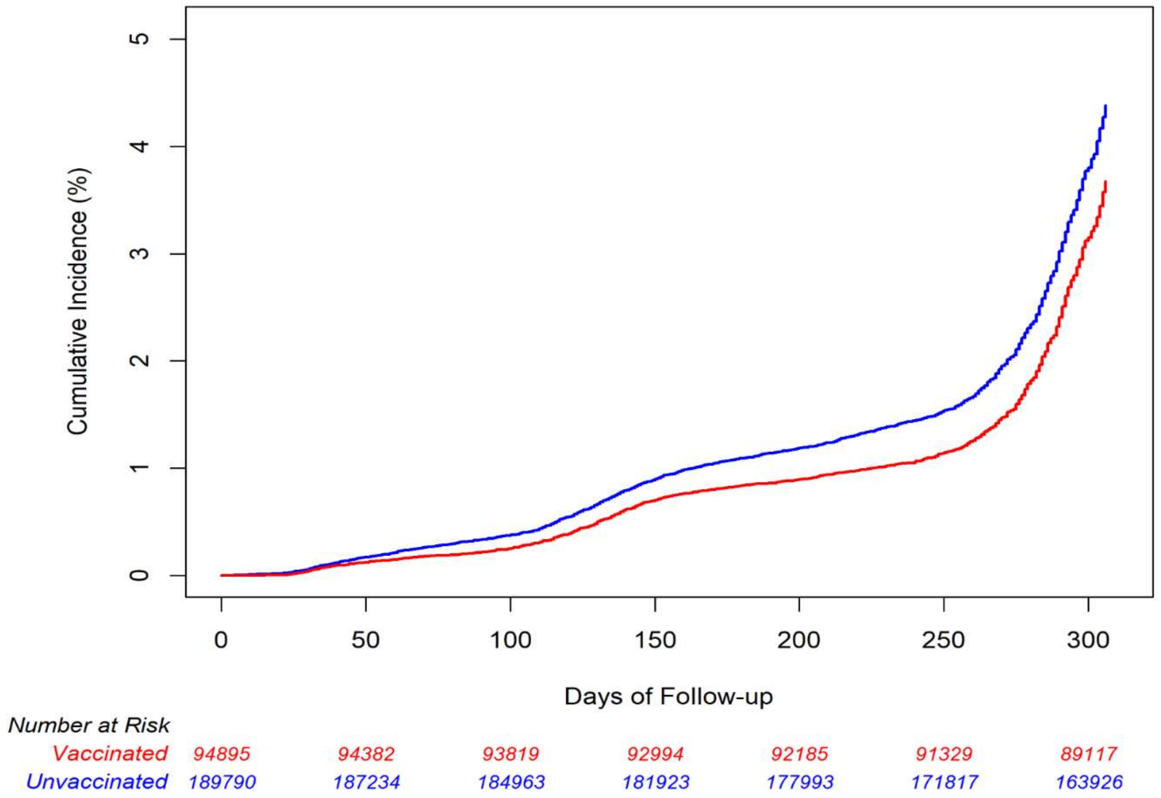


Abbreviations: COVID-19, coronavirus disease 2019; RZV, recombinant zoster vaccine

**Supplementary Figure 2: Cumulative incidence estimates of COVID-19 hospitalization by RZV (2 dose) vaccination status**

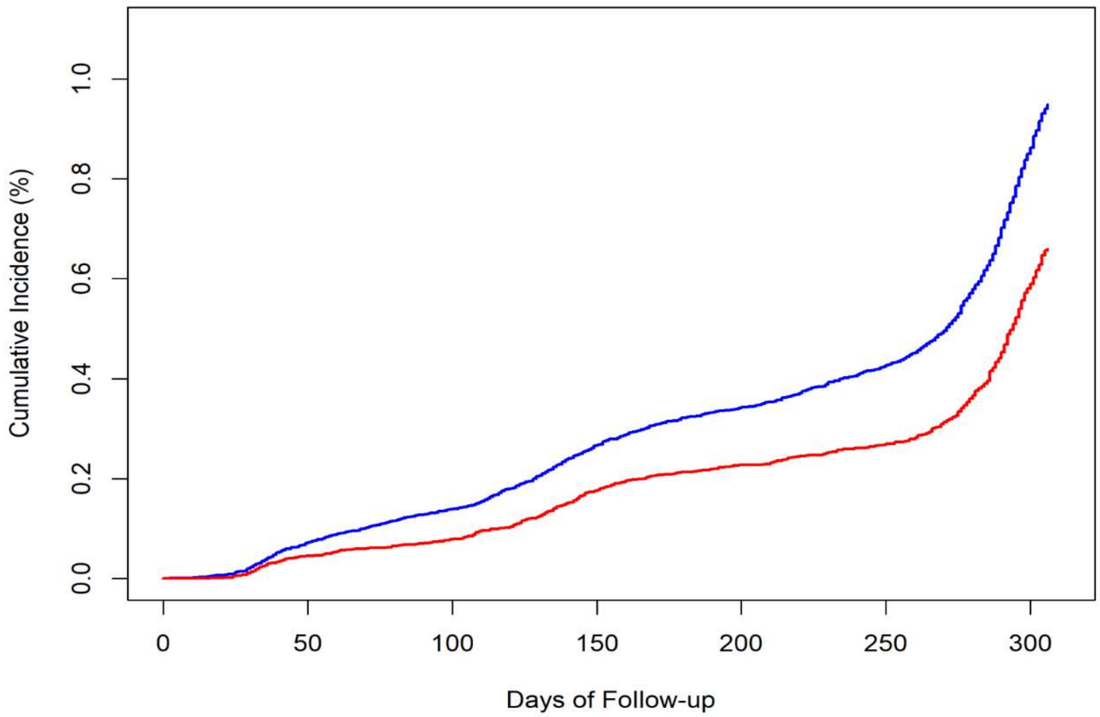


| *Number at Risk* | | | | | | | |
|---|---|---|---|---|---|---|---|
| *Vaccinated* | *94895* | *94449* | *93975* | *93478* | *92791* | *92115* | *91434* |
| *Unvaccinated* | *189790* | *187392* | *185346* | *182987* | *179387* | *173552* | *168671* |

Abbreviations: COVID-19, coronavirus disease 2019; RZV, recombinant zoster vaccine
